# Association Between Abnormal Sleep and Mortality Risk: A Nationwide Prospective Cohort Study of All-Cause and Cardiovascular disease Mortality

**DOI:** 10.1101/2025.02.21.25322660

**Authors:** Li Ke, Ying Li, Lei Zhao, Wenli Xing, Sili Jiang

**Affiliations:** Department of Cerebrovascular Diseases, Suining Central Hospital, Suining, Sichuan Province, China

**Keywords:** Sleep duration, All-cause mortality, CVD mortality, National Health and Nutrition Examination Survey

## Abstract

**Background:** The American Heart Association’s Life’s Essential 8 construct of ideal cardiovascular health now includes sleep duration, emphasizing the need to understand the implications of suboptimal sleep patterns. Recent studies have demonstrated that Life’s Essential 8 is correlated with the risk of all-cause mortality and CVD-specific mortality. However, although sleep duration being one of the components of Life’s Essential 8, the relationship between an individual’s sleep patterns and the risk of all-cause and CVD- specific mortality remains unclear.

**Methods:** The 2005–2018 National Health and Nutrition Examination Survey (NHANES) included 38,006 adults aged ≥20 years. The definitions of normal sleep duration (7–8 hours per night), short sleep duration (< 7 hours), and long sleep duration (> 8 hours) were based on the current literature. To evaluate the association between abnormal sleep patterns and both all-cause mortality and CVD mortality, a multinomial Cox regression analysis was conducted, adjusting for demographic and clinical factors.

**Results:** The study utilized three models (Model I–III), first unadjusted and then adjusted for sociodemographic, lifestyle, and chronic disease factors. In the fully adjusted model, both short sleep (OR=1.25, 95% CI: 1.12–1.39, P<0.001) and long sleep (OR=1.30, 95% CI: 1.18–1.44, P<0.001) were found to be associated with an increased risk of all-cause mortality. Regarding CVD mortality, after full adjustment (Model III), only long sleep remained statistically significant (OR=1.28, 95% CI: 1.07–1.52, P=0.006), while the association for short sleep was no longer significant (OR=1.16, 95% CI: 0.94–1.43, P=0.176).

**Conclusions:** Abnormal sleep duration (both short and long) was independently associated with an increased risk of all-cause mortality. Long sleep duration was associated with an increased risk of CVD mortality.

## Introduction

Sleep behavior is a crucial indicator of physiological health. Numerous epidemiological studies have investigated the relationship between sleep duration and various health-related outcomes[1–4]. Sleep significantly influences the human autonomic nervous system, vascular tone, cardiac performance, endothelial activity, and coagulation[5,6]. Previous researches has suggested that adults should aim for 7–8 hours of sleep per night[7,8]. Furthermore, studies indicate that both short and long sleep durations can adversely affect hormonal levels, metabolism, and immune function[7,9,10]. The American Heart Association’s Life’s Essential 8 construct of ideal cardiovascular health now includes sleep duration[11], emphasizing the need to understand the implications of suboptimal sleep patterns. Recent studies have demonstrated that Life’s Essential 8 is correlated with the risk of all-cause mortality and cardiovascular disease (CVD)-specific mortality[12]. However, although sleep duration being one of the components of Life’s Essential 8, the relationship between an individual’s sleep patterns and the risk of all-cause and CVD-specific mortality remains unclear. We examined associations between sleep duration and all-cause/cardiovascular disease (CVD) mortality using 2005–2018 NHANES data from 38,006 nationally representative U.S. adults (aged ≥20 years).

## Methods

### Study population

The National Health and Nutrition Examination Surveys (NHANES) is a nationally representative study designed to evaluate the health and nutritional status of the non-institutionalized U.S. population. The NHANES protocol underwent comprehensive review and received approval from the National Center for Health Statistics Research Ethics Review Board, with informed consent obtained from all participants. The institutional review board of Suining Central Hospital classified the study as exempt due to the use of publicly available and deidentified data, thereby waiving the need for informed consent. This research adhered to the Strengthening the Reporting of Observational Studies in Epidemiology (STROBE) guidelines to ensure adherence to quality reporting standards. Our study used the NHANES data from the 7 consecutive NHANES data cycles spanning 2005 to 2018. Data from adult participants (aged 20 years and older) were collected. Participants lacking information on sleep duration and those who were lost to follow-up were excluded. Ultimately, a total of 38,006 participants were included in the analysis (Figure. 1).

**Figure 1.**
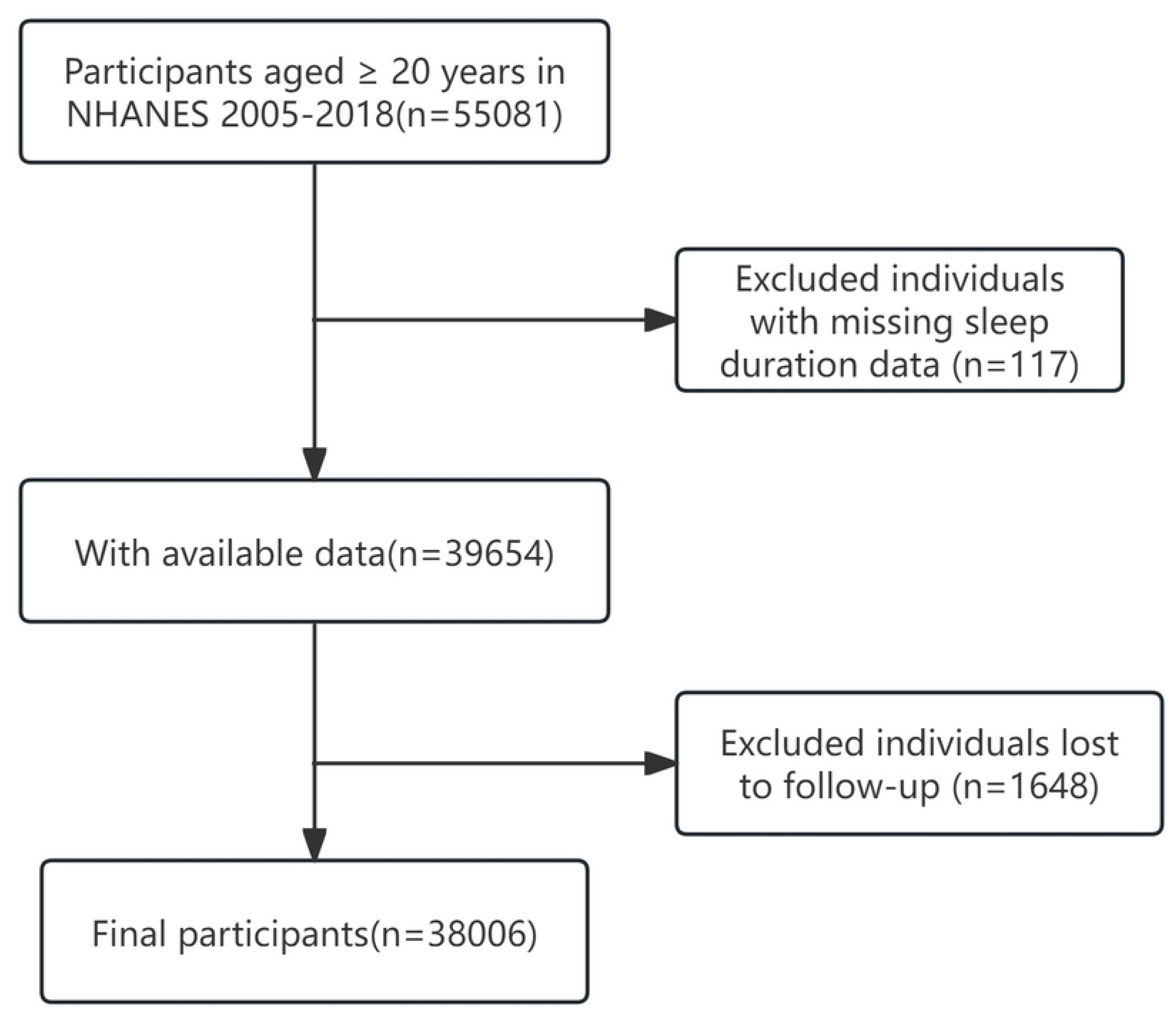
Study flow chart.

### Assessment of Sleep Duration

In the seven NHANES cycles conducted from 2005 to 2018, sleep duration was assessed using the question, “How much sleep do you usually get at night on weekdays or workdays?” Participants reported their typical daily sleep hours. Sleep duration was categorized into three commonly referenced groups[3,4]: Short sleep (<7 hours), normal sleep (7–8 hours), and long sleep (≥8 hours).

### Assessment of mortality

Data on mortality were obtained by linking the cohort database to the National Death Index from the Centers for Disease Control, with the reference date set to 31 December 2019. The cardiovascular mortality included in this analysis covers a variety of ICD codes, specifically: I00–I09 for acute and chronic rheumatic heart diseases; I11 for hypertensive heart disease; I13 for hypertensive heart disease with renal involvement; I20– I25 for ischemic heart diseases; I26–I28 for pulmonary embolism and other acute pulmonary heart conditions; I29 for other cardiovascular diseases; I30–I51 for other heart diseases; and I60–I69 for cerebrovascular diseases.

### Assessment of covariates

Covariates were selected through a hybrid approach incorporating variables with univariate associations (P < 0.05) and clinically relevant factors (P ≥ 0.05). Additional confounders were included if they demonstrated ≥10% impact on effect estimates or plausible biological links to outcomes. The analysis adjusted for Age, sex, race/ethnicity, marital status, education level, poverty-to-income ratio (PIR), body mass index (BMI), smoking and alcohol status, and medical history, including conditions like hypertension, hyperlipidaemia, diabetes. Race/ethnicity included non-Hispanic White, non-Hispanic Black, Mexican American, and other groups. Marital status comprised married, living with a partner, or living alone. Education level was stratified as <9, 9–12, or >12 years. Family income was categorized by poverty-to-income ratio (PIR) as low (≤1.3), medium (1.3–3.5), or high (>3.5) following U.S. federal guidelines. Smoking status was categorized as: never: <100 lifetime cigarettes; former: ≥100 cigarettes with no current use; and current: ≥100 cigarettes with occasional or daily use. Alcohol use was self-reported and defined as: never: <12 lifetime drinks; former: ≥12 drinks/year historically, none in the past year; Current: ≥12 drinks/year with recent consumption. Physical activity (PA) is defined as the duration that individuals dedicate to participating in various activities, including walking, cycling, performing household tasks, engaging in work-related duties and pursuing recreational activities throughout the week. If no exercise occurs within a week, the recorded exercise time is considered to be zero. Body mass index (BMI) was calculated as weight/height² (kg/m²).

Hyperlipidaemia is defined by one or more of the following conditions: the administration of lipid-lowering drugs; increased triglyceride levels (≥150 mg/dL); or heightened cholesterol levels, which encompass total cholesterol (≥200 mg/dL), LDL cholesterol (≥130 mg/dL), or HDL cholesterol (<40 mg/dL). Hypertension and diabetes status were confirmed through self-reported physician diagnoses.

### Statistical Analysis

Analyses adhered to NHANES protocols, incorporating sampling weights and design variables to account for complex survey structure. Data were derived from family interviews and Mobile Examination Center (MEC) assessments, so the weights provided by MECs should be utilised. Given the low percentage of missing data (0–9% across variables), a multivariate single imputation technique was utilised. Continuous variables are presented as weighted means (SD) or medians; categorical variables as weighted counts (%). Group differences were assessed using weighted ANOVA (continuous) and chi-square tests (categorical). Weighted multivariable COX regression models estimated odds ratios (HR) with 95% confidence intervals (CI) for associations between sleep duration and all-cause/cardiovascular mortality. Covariates were hierarchically adjusted across models: Model 1: Demographic factors (age, sex, race), socioeconomic status (education, poverty income ratio [PIR]), marital status, and NHANES cycle. Model 2: Model 1 + lifestyle factors (physical activity, BMI, alcohol consumption, smoking status). Model 3: Model 2 + clinical comorbidities (hypertension, hyperlipidemia, diabetes).

Furthermore, we conducted subgroup analyses based on sex (men vs. women), age (<60 vs. ≥60 years), race/ethnicity (non-Hispanic White vs. other), BMI (<30 vs. ≥30 kg/m²), smoking status (never vs. former or current), alcohol consumption status (never vs. former or current), hypertension (no vs. yes), and diabetes (no vs. yes) utilizing multivariable Cox proportional hazards regression models. All analyses were conducted by utilising the statistical software packages R4.2.2 (available at http://www.R-project.org) and Free Statistics software version 2.1 (Beijing Free Clinical Medical Technology Co., Ltd.^2^).

## Results

### Baseline characteristics

Table 1 presents the weighted demographics and health characteristics of participants categorized by sleep duration (short: <7 hours; normal: 7-8 hours; long: >8 hours) in the NHANES 2005-2018. In the overall sample, the proportion of males was higher in the short sleep group (51.5%) than in the normal sleep group (48.3%) or the long sleep group (43.5%). Conversely, females constituted 56.5% of the long sleep group. Additionally, the age distribution shows that the average age of participants in the long sleep group (48.40 years) is slightly higher than the overall average (47.37 years). Most participants in all groups were non- Hispanic white and had received high school education or higher.

**Tabel 1.**
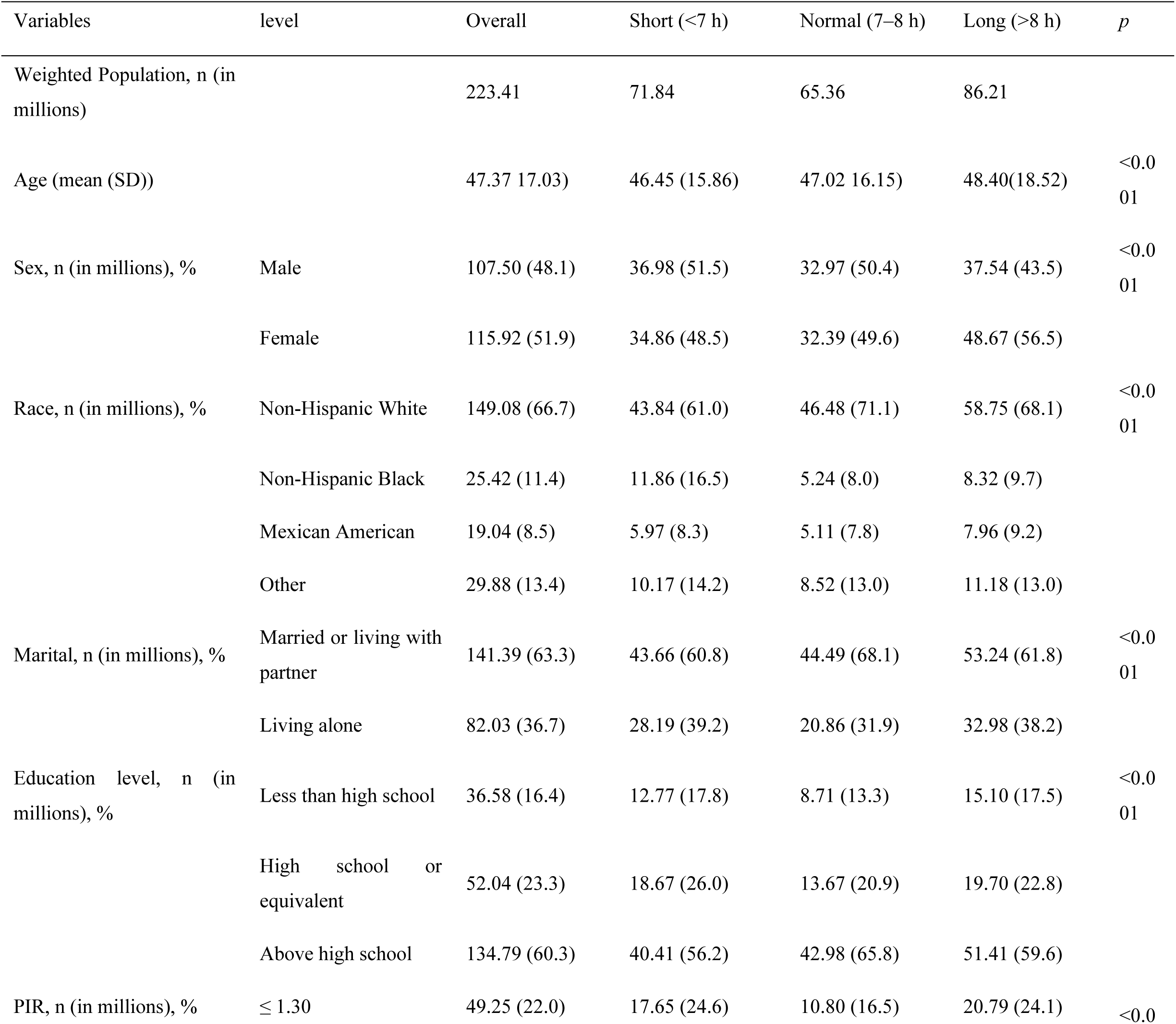
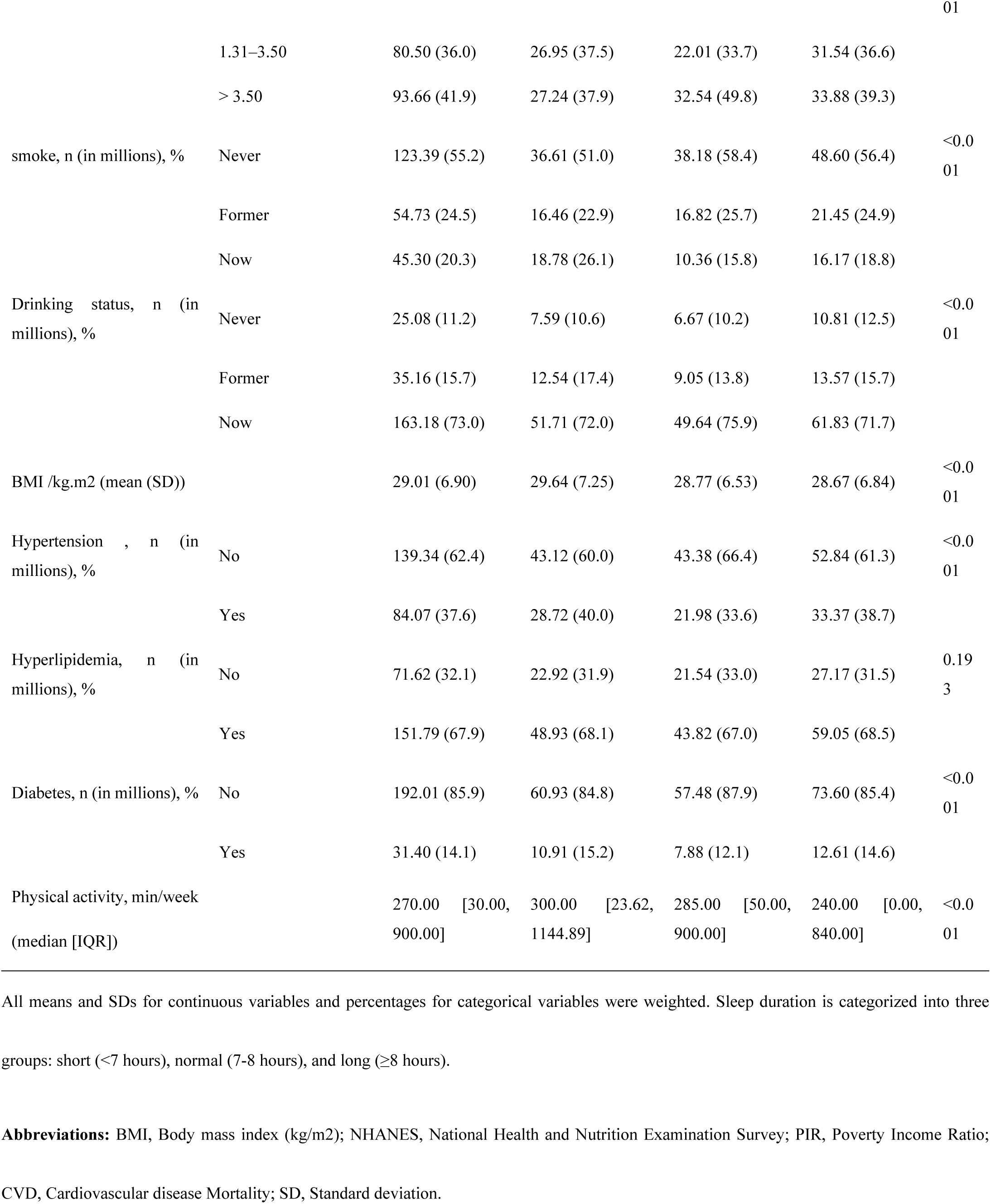
Demographics of Sleep Duration in NHANES 2005–2018.

### Abnormal sleep and mortality

During 1–14 years of follow-up in NHANES 2005–2018, a total of 4,200 all-cause deaths and 1,262 CVD deaths were identified. Table 2 shows the relationship between sleep duration and the risk of all-cause and CVD mortality, as assessed using a multifactorial Cox regression model. The study used three models (Model I–III), which were initially unadjusted and then adjusted for sociodemographic, lifestyle, and chronic disease factors. In the unadjusted model, both short sleep (<7 hours; OR=1.42, 95% CI: 1.28–1.58, P<0.001) and long sleep (≥8 hours; OR=1.83, 95% CI: 1.63–2.05, P<0.001) were associated with an increased risk of all-cause mortality. After full adjustment (Model III), the risks remained significant for short sleep (OR=1.25, 95% CI: 1.12–1.39, P<0.001) and long sleep (OR=1.30, 95% CI: 1.18–1.44, P<0.001), with a significant dose-response trend observed (P for trend<0.001). In the unadjusted model, both short sleep (OR=1.35, 95% CI: 1.09–1.67, p=0.005) and long sleep (OR=1.95, 95% CI: 1.63–2.35, p<0.001) were associated with increased risks of CVD mortality. However, after full adjustment (Model III), only long sleep remained statistically significant (OR=1.28, 95% CI: 1.07–1.52, p=0.006), while short sleep was no longer significant (OR=1.16, 95% CI: 0.94–1.43, p=0.176). A significant trend for long sleep was observed (p for trend=0.005).

**Tabel 2.**
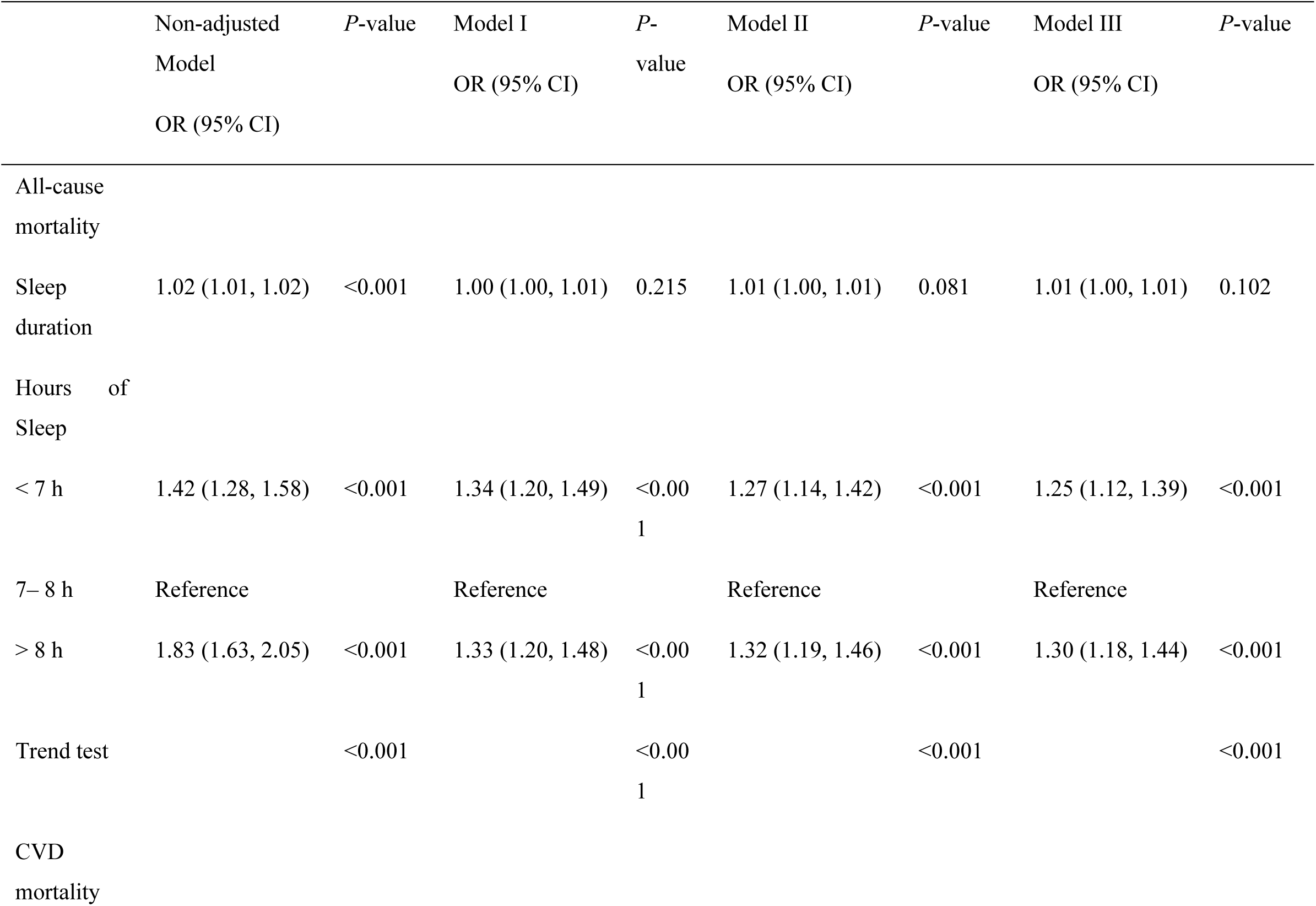
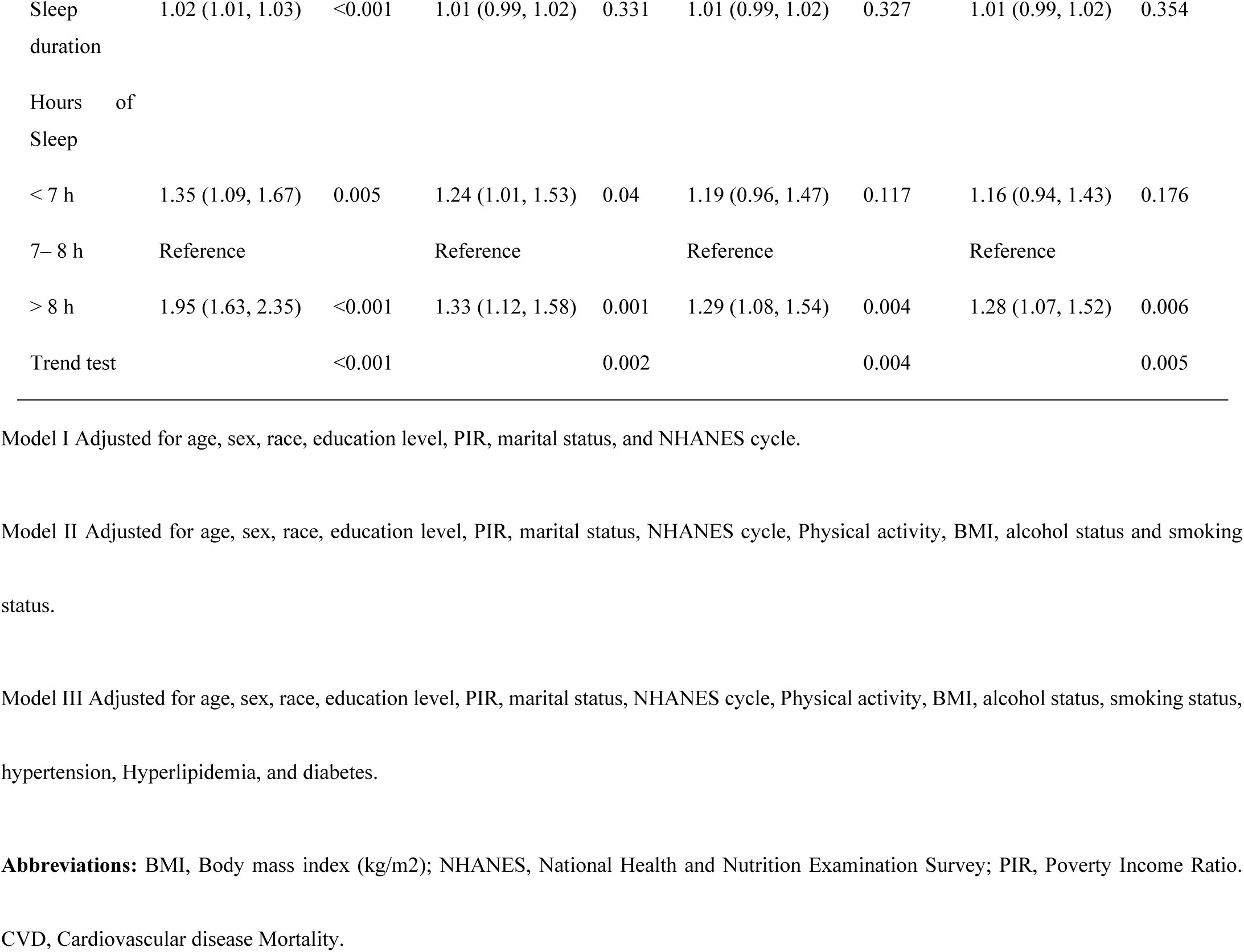
Multinomial Cox Regression Analysis of Short and Long Sleep Durations in Individuals at Risk of All-Cause and CVD Mortality.

|  | Non-adjusted<br>Model<br>OR (95% CI) | P-value | Model I<br>OR (95% CI) | P-value | Model II<br>OR (95% CI) | P-value | Model III<br>OR (95% CI) | P-value |
| --- | --- | --- | --- | --- | --- | --- | --- | --- |
| All-cause<br>mortality |  |  |  |  |  |  |  |  |
| Sleep<br>duration | 1.02 (1.01, 1.02) | <0.001 | 1.00 (1.00, 1.01) | 0.215 | 1.01 (1.00, 1.01) | 0.081 | 1.01 (1.00, 1.01) | 0.102 |
| Hours of<br>Sleep |  |  |  |  |  |  |  |  |
| < 7 h | 1.42 (1.28, 1.58) | <0.001 | 1.34 (1.20, 1.49) | <0.001 | 1.27 (1.14, 1.42) | <0.001 | 1.25 (1.12, 1.39) | <0.001 |
| 7– 8 h | Reference |  | Reference |  | Reference |  | Reference |  |
| > 8 h | 1.83 (1.63, 2.05) | <0.001 | 1.33 (1.20, 1.48) | <0.001 | 1.32 (1.19, 1.46) | <0.001 | 1.30 (1.18, 1.44) | <0.001 |
| Trend test |  | <0.001 |  | <0.001 |  | <0.001 |  | <0.001 |
| CVD<br>mortality |  |  |  |  |  |  |  |  |
| Sleep duration | 1.02 (1.01, 1.03) | <0.001 | 1.01 (0.99, 1.02) | 0.331 | 1.01 (0.99, 1.02) | 0.327 | 1.01 (0.99, 1.02) | 0.354 |
| Hours of Sleep |  |  |  |  |  |  |  |  |
| < 7 h | 1.35 (1.09, 1.67) | 0.005 | 1.24 (1.01, 1.53) | 0.04 | 1.19 (0.96, 1.47) | 0.117 | 1.16 (0.94, 1.43) | 0.176 |
| 7– 8 h | Reference |  | Reference |  | Reference |  | Reference |  |
| > 8 h | 1.95 (1.63, 2.35) | <0.001 | 1.33 (1.12, 1.58) | 0.001 | 1.29 (1.08, 1.54) | 0.004 | 1.28 (1.07, 1.52) | 0.006 |
| Trend test |  | <0.001 |  | 0.002 |  | 0.004 |  | 0.005 |

Model I Adjusted for age, sex, race, education level, PIR, marital status, and NHANES cycle.
Model II Adjusted for age, sex, race, education level, PIR, marital status, NHANES cycle, Physical activity, BMI, alcohol status and smoking status.
Model III Adjusted for age, sex, race, education level, PIR, marital status, NHANES cycle, Physical activity, BMI, alcohol status, smoking status, hypertension, Hyperlipidemia, and diabetes.
**Abbreviations:** BMI, Body mass index (kg/m2); NHANES, National Health and Nutrition Examination Survey; PIR, Poverty Income Ratio. CVD, Cardiovascular disease Mortality.

The Kaplan-Meier curves in Figure 2 show All-cause and CVD mortality, comparing the survival rates of the short and long sleep cohorts to those of the normal sleep group. Notably, survival rates were significantly lower in both short and long sleep cohorts compared to the normal sleep group.

**Figure 2.**
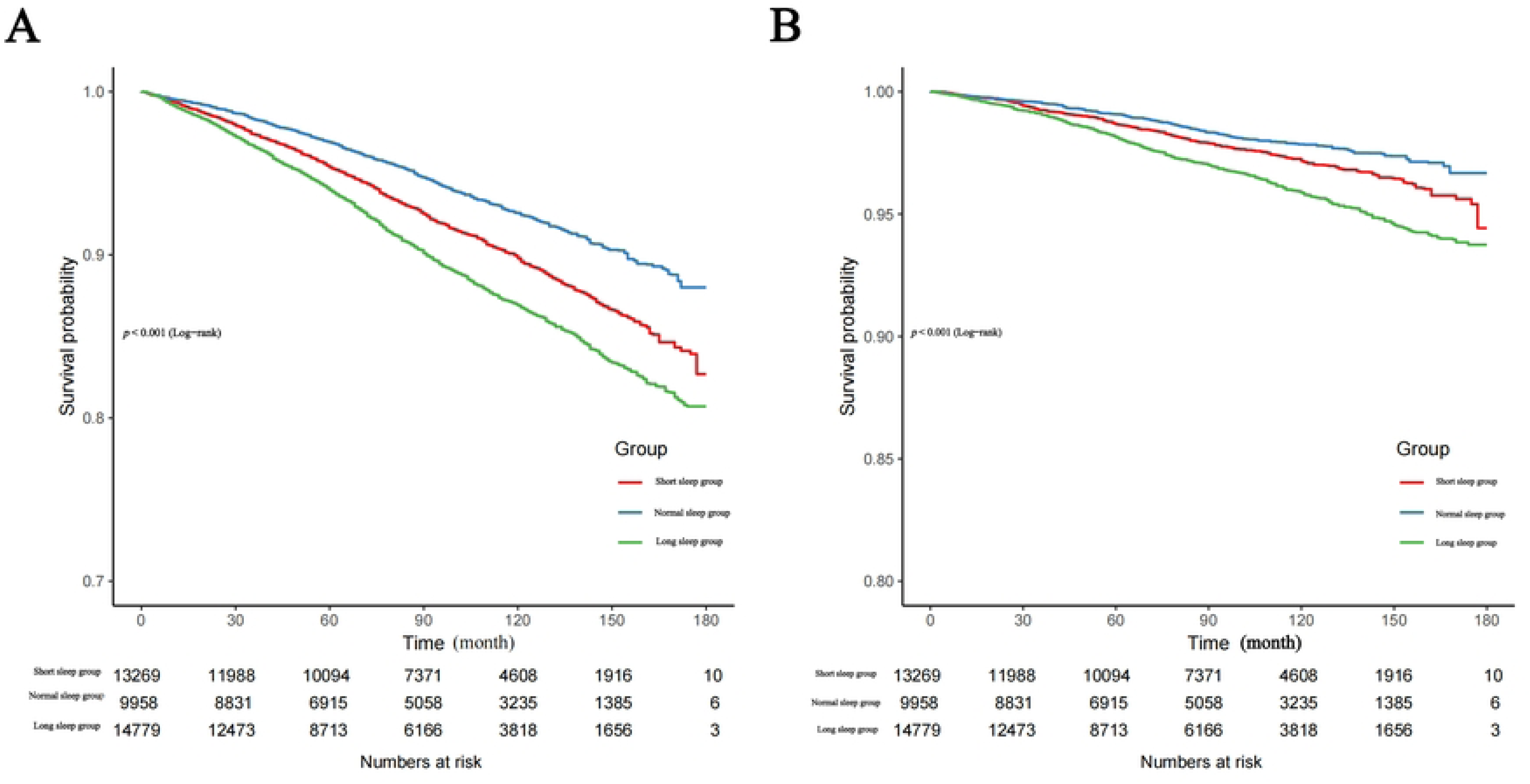
A: Kaplan-Meier survival curve for all-cause mortality among individuals with short, long, and normal sleep. B: Kaplan-Meier survival curve for CVD mortality among individuals with short, long, and normal sleep. **Abbreviations:** CVD, Cardiovascular disease Mortality.

### Subgroup analyses

Figure 3 presents subgroup analyses examining the association between sleep duration and both All-cause and CVD mortality. Compared to normal sleep, we found that age and race influenced the association between sleep duration (short or long) and all-cause mortality (P < 0.05 for interaction). However, we did not observe any significant interactions among other subgroups. In stratified analyses by sex (men and women), hypertension status (with and without), and diabetes status (without diabetes), compared with normal sleep, the risk of all-cause mortality in the short and long sleep groups showed a stronger association. In the subgroup analysis examining the relationship between sleep duration and CVD mortality, after adjusting for multiple factors (Model III), prolonged sleep (> 8 hours) was associated with a significant risk in certain subgroups. In contrast, the association with short sleep (< 7 hours) was weak, and no significant interactions were detected across any subgroup.

**Figure 3.**
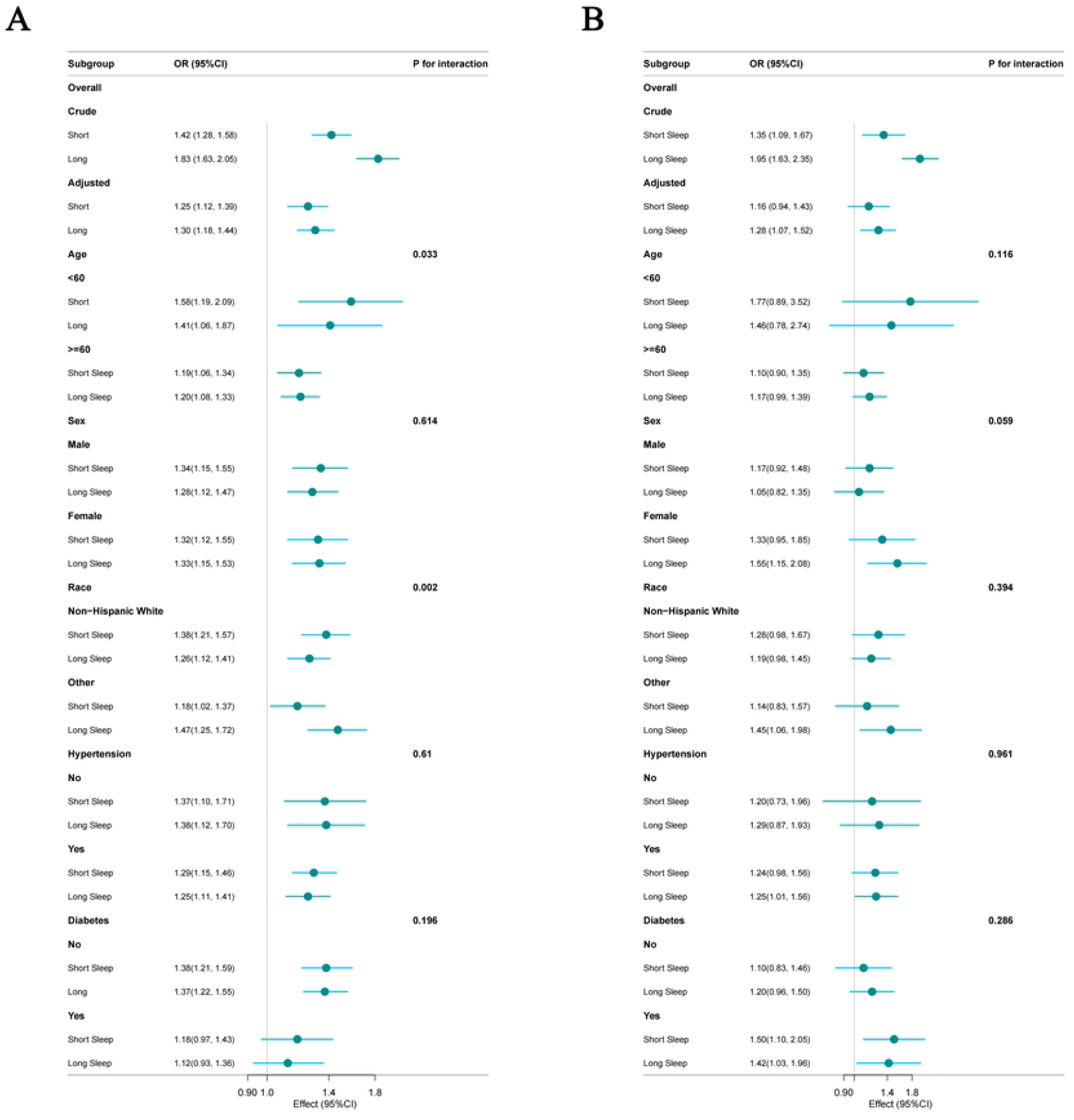
A: The association of short and long sleep with all-cause mortality was analyzed in relation to general characteristics, the normal sleep group was used as the reference group. B: The association of short and long sleep with CVD mortality was analyzed in relation to general characteristics, the normal sleep group was used as the reference group. The stratifications were adjusted for age, sex, race, education level, PIR, marital status, NHANES cycle, Physical activity, BMI, alcohol status, smoking status, hypertension, Hyperlipidemia, and diabetes. **Abbreviations:** BMI, Body mass index (kg/m2); NHANES, National Health and Nutrition Examination Survey; PIR, Poverty Income Ratio. CVD, Cardiovascular disease Mortality.

### Sensitivity analysis

The results of the sensitivity analyses are presented in Table 3. After excluding participants who died within 2 years of follow-up and those with a history of cancer at baseline, the adjusted hazard ratios (HR) for all-cause mortality were 1.24 (95% CI: 1.06–1.45; P < 0.001) for short sleep and 1.33 (95% CI: 1.16–1.52; P < 0.001) for long sleep, compared to normal sleep. For CVD mortality, the adjusted HR was 1.23 (95% CI: 0.95–1.61; P=0.118) for short sleep and 1.35 (95% CI: 1.12–1.63; P=0.002) for long sleep (>8 hours).

**Tabel 3.** Sensitivity analyses, excluding those with cancer and died within 2 years of follow-up.

|  | Non-adjusted<br>Model<br>OR (95% CI) | *P*-value | Model I<br>OR (95% CI) | *P*-<br>value | Model II<br>OR (95% CI) | *P*-value | Model III<br>OR (95% CI) | *P*-<br>value |
| --- | --- | --- | --- | --- | --- | --- | --- | --- |
| All-cause mortality |  |  |  |  |  |  |  |  |
| Sleep duration | 1.01 (1.01, 1.02) | <0.001 | 1.00 (0.99, 1.01) | 0.582 | 1.00 (0.99, 1.01) | 0.963 | 1.00 (0.99, 1.01) | 0.896 |
| Hours of Sleep |  |  |  |  |  |  |  |  |
| < 7 h | 1.47 (1.28, 1.69) | <0.001 | 1.33 (1.15, 1.55) | <0.001 | 1.27 (1.08, 1.49) | 0.003 | 1.24 (1.06, 1.45) | 0.007 |
| 7– 8 h | Reference |  | Reference |  | Reference |  | Reference |  |
| > 8 h | 1.83 (1.58, 2.11) | <0.001 | 1.36 (1.18, 1.57) | <0.001 | 1.35 (1.18, 1.55) | <0.001 | 1.33 (1.16, 1.52) | <0.001 |
| Trend test |  | <0.001 |  | <0.001 |  | <0.001 |  | <0.001 |
| CVD mortality |  |  |  |  |  |  |  |  |
| Sleep duration | 1.01 (1.00, 1.02) | 0.003 | 0.99 (0.98, 1.01) | 0.462 | 0.99 (0.98, 1.01) | 0.415 | 0.99 (0.98, 1.01) | 0.391 |
| Hours of Sleep |  |  |  |  |  |  |  |  |
| < 7 | 1.50 (1.17, 1.91) | 0.001 | 1.32 (1.02, 1.72) | 0.037 | 1.26 (0.96, 1.65) | 0.091 | 1.23 (0.95, 1.61) | 0.118 |
| 7– 8 | Reference |  | Reference |  | Reference |  | Reference |  |
| ≥ 8 | 2.04 (1.67, 2.49) | <0.001 | 1.41 (1.15, 1.72) | <0.001 | 1.37 (1.12, 1.66) | 0.002 | 1.35 (1.12, 1.63) | 0.002 |
| Trend test |  | <0.001 |  | 0.001 |  | 0.002 |  | 0.001 |

- 184 - Model I Adjusted for age, sex, race, education level, PIR, marital status, and NHANES cycle. - 185 - Model II Adjusted for age, sex, race, education level, PIR, marital status, NHANES cycle, Physical activity, BMI, alcohol status and smoking - 186 - status. - 187 - Model III Adjusted for age, sex, race, education level, PIR, marital status, NHANES cycle, Physical activity, BMI, alcohol status, smoking - 188 - status, hypertension, Hyperlipidemia, and diabetes. - 189 - Abbreviations:** BMI, Body mass index (kg/m2); NHANES, National Health and Nutrition Examination Survey; PIR, Poverty Income Ratio. - 190 - CVD, Cardiovascular disease Mortality.

## Discussion

In this nationally representative study, we identified a robust association between sleep duration and all- cause and CVD mortality. Our comparison of individuals with long and short sleep durations versus those with normal sleep revealed that both long and short sleepers exhibited increased risk of all-cause mortality. Moreover, individuals reporting long sleep duration had an elevated risk of death attributable to CVD mortality. Subgroup analysis indicated that age and race interactively influence all-cause mortality, with age potentially exacerbating these associations. We posit that future large-scale, meticulously designed cohort studies are crucial for achieving precise estimates.

In a previous meta-analysis of 27 cohort studies conducted by Da Silva et al., both long and short sleep durations were significantly associated with an increased risk of all-cause mortality among older adults[13]. Our findings indicate that the risk of all-cause mortality associated with both short and long sleep is significantly increased in middle-aged and younger individuals (under 60 years of age). The potential reason is that we included middle-aged and younger individuals and adjusted for various confounding factors.

Several studies have shown that, compared to individuals sleeping 7-8 hours nightly, those sleeping longer or shorter durations face a greater risk of adverse outcomes[14]. Both insufficient and excessive sleep are linked to overall mortality[15], type 2 diabetes[16], hypertension[17,18], respiratory diseases[19], adult obesity[20], and self-assessed poor health[21]. A Japanese study involving individuals with type 2 diabetes indicates that those with insufficient or prolonged sleep are at a greater risk of developing metabolic syndrome and insulin resistance[22]. Furthermore, a significant association has been observed between sleep duration and breast cancer risk, women who slept less than or equal to 6.0 hours per day (OR (95% CI): 1.53 (1.10-12.12)) and those who slept longer than 8.9 hours per day (OR (95% CI): 1.59 (1.17-20.17)) had an increased risk of breast cancer[23]. These clinical studies support that short and long sleep durations are significantly associated with an increased risk of all-cause mortality.

A systematic review has revealed that prolonged sleep duration is associated with higher mortality rates and an increased incidence of various health conditions[24,25]. The relative risks (RRs) identified include stroke (1.46), CVD (1.25), and coronary heart disease (1.24) [24]. Furthermore, the prevalence of depression correlates with variations in sleep duration, as studies indicate that when sleep duration exceeds 8 hours, the risk of depression significantly increases with longer sleep durations[3]. Researches has shown that depression has a substantial negative impact on the development of CVD and on CVD mortality[26–28].

The mechanisms linking sleep to all-cause mortality and CVD death remain unclear. One proposed mechanism connecting short sleep to mortality involves the association between short sleep and decreased leptin and increased ghrelin (a peptide that stimulates appetite) [29]. This heightened appetite may elucidate the connection between sleep deprivation and obesity [30], as well as the onset of diabetes[31]. Conversely, the association between obesity, diabetes, and mortality outcomes are well established. Additionally, longer sleep duration is associated with increased plasma levels of IL-10, CRP, and IL-6[19,32]. Inflammation is recognized as a risk factor for neurodegenerative diseases[33] and contributes to a higher incidence rate of CVD[34].

This study examined the relationship between sleep duration and all-cause and cardiovascular disease (CVD) mortality using data from the National Health and Nutrition Examination Survey (NHANES). The sample size is large and representative, which facilitates a comprehensive analysis. However, this study has some limitations. The determination of sleep duration relies on participants’ self-reports, and the various reasons for sleep deprivation may be associated with different demographic or personality traits, such as low socioeconomic status or high neuroticism. Further research is needed to explore these subtle factors to develop more relevant and effective sleep health interventions.

## Conclusions

In summary, our research utilizing data from the NHANES, covering the period from 2005 to 2018, indicates that sleep duration is associated with both all-cause mortality and cardiovascular mortality. Specifically, both short and long sleep durations are linked to an increased risk of all-cause mortality, whereas prolonged sleep duration is associated with a heightened risk of CVD mortality.

## Data availability statement

The public datasets utilized in this study are available online. Information regarding the names of the repositories and their corresponding accession numbers can be found at https://www.cdc.gov/nchs/nhanes/. It is important to note that no new datasets were generated or analyzed in the present study.

## Ethics statement

The protocol for the NHANES study received approval from the Research Ethics Review Committee of the National Center for Health Statistics (NCHS). All participants provided written informed consent. The institutional review board of Suining Central Hospital deemed the study exempt due to its use of publicly accessible and de-identified data, thereby waiving the requirement for informed consent.

## Author contributions

Study concept and design: Li Ke, Ying Li, Wenli Xing, Lei Zhao and Sili Jiang. Acquisition of data: Li Ke, Ying Li. Analysis and interpretation of data: Li Ke, Sili Jiang and Wenli Xing. Wrote of themanuscript: Li Ke, Lei Zhao, Sili Jiang, Wenli Xing, and Ying Li. All authors have revised the manuscript.

## Acknowledgments

The authors would like to acknowledge the NHANES staff, researchers, and participants for their contributions. Additionally, we wish to thank Dr. Jie Liu from the People’s Liberation Army General Hospital in Beijing, China, for his support with statistical analyses, advice on study design.

